# Health Literacy and Cardiovascular Health Among Formerly Incarcerated Individuals: Evidence from the JUSTICE Study

**DOI:** 10.64898/2026.10.23.26366373

**Authors:** Bahar N. Bouzarjomehri, Lisa B. Puglisi, Emily A. Wang, Jenerius A. Aminawung

**Author notes:** Corresponding Author: Jenerius A. Aminawung, MD, MPH SEICHE Center for Health and Justice 60 Temple St, Suite 5C, New Haven, CT 06510.

## Abstract

**Background:** Health literacy, defined as the ability to find, understand, and use health information, is a key determinant of cardiovascular disease but remains underexamined among individuals transitioning from incarceration to community life. This study examined the association between health literacy and cardiovascular health among adults recently released from incarceration.

**Methods:** Using baseline data from the Justice-Involved Individuals Cardiovascular Disease Epidemiology (JUSTICE) study, a prospective cohort of individuals with cardiovascular risk factors released from correctional facilities, we examined the association between health literacy, as measured by the Short Test of Functional Health Literacy in Adults, and cardiovascular health, as measured by the American Heart Association’s Life’s Essential 8 (LE8).

**Results:** Among 439 participants (mean age 45 years), 32 (7%) demonstrated inadequate health literacy. Hispanic ethnicity (53% vs. 25%), less than high school education (50% vs. 20%), and homelessness (16% vs. 3.2%) were significantly associated with inadequate health literacy. In bivariate analyses, inadequate health literacy was associated with lower LE8 blood glucose (63.1 vs. 76.8, p = 0.02), and LE8 physical activity (19.4 vs. 32.2, p = 0.03), but higher LE8 diet (55.5 vs. 43.8, p = 0.04) scores. After adjusting for age, gender, race/ethnicity, education level, and housing status, inadequate health literacy remained independently associated with lower LE8 glucose scores (β = −11.9, 95% CI: −22.9 to −0.8, p = 0.04).

**Conclusion:** Health literacy is a modifiable determinant of cardiovascular health, particularly glycemic control, in justice-involved populations. Interventions strengthening health literacy may help reduce health disparities among formerly incarcerated individuals.

**Clinical Perspective:** *What is new?:* We find that lower health literacy among individuals recently released from incarceration is associated with poorer glycemic control, a key component of cardiovascular health, as measured by the American Heart Association’s Life’s Essential 8 framework.

*What are the clinical implications?:* Our data suggest that health literacy may represent a modifiable factor in cardiovascular risk reduction among formerly incarcerated individuals. Integrating literacy-sensitive education, chronic disease self-management support, and improved care coordination between the carceral and community health systems could strengthen cardiovascular prevention and promote long-term health equity.

## Introduction

According to the U.S. Department of Health and Human Services, health literacy is defined as “the degree to which individuals have the ability to find, understand, and use information and services to inform health-related decisions and actions for themselves and others” [1]. Limited health literacy is increasingly recognized as a social determinant of health and an independent predictor of morbidity, healthcare utilization, and mortality [1–3]. Inadequate health literacy is disproportionately prevalent among populations experiencing structural disadvantage, including those who are formerly incarcerated [2, 4–6]. Factors strongly associated with low health literacy, such as low educational attainment, housing instability, and low income, are often linked to and compounded by incarceration [1, 3, 7].

The transition from incarceration to community further exacerbates health vulnerabilities, as many individuals return to the community with newly diagnosed chronic conditions, fragmented care continuity, and limited insurance coverage [6, 8, 9]. For instance, many who are recently released and living in halfway houses do not have access to cell phones or the internet, limiting their ability to obtain health information. Consequently, formerly incarcerated individuals frequently face steep barriers in navigating healthcare systems and independently managing chronic illness [6].

These challenges are especially consequential in the context of cardiovascular disease (CVD), a leading cause of morbidity and mortality in justice-impacted communities. An estimated 10% of the 1.8 million incarcerated individuals in the United States are diagnosed with CVD [5, 10, 11]. Effective CVD management relies heavily on patients’ ability to understand treatment instructions, manage medications, monitor blood pressure and glucose, and sustain dietary and physical activity goals [12, 13]. Individuals with inadequate health literacy are more likely to struggle with this self-management and depend more heavily on emergency care, underscoring health literacy’s role as a key social determinant of cardiovascular health, as identified by the American Heart Association [14, 15].

Despite the substantial burden of CVD in incarcerated communities, little is known about the relationship between health literacy and cardiovascular health in this population. A recent study of incarcerated individuals identified a high prevalence of inadequate health literacy associated with increased cardiovascular risk [5], but no studies to our knowledge have characterized this relationship among those recently released. The objective of this study was to examine the association between health literacy and cardiovascular health among individuals recently released from incarceration. By focusing on individuals during the transition from incarceration to the community, this study aims to address a critical gap in understanding how modifiable social determinants, such as health literacy, contribute to cardiovascular risk in justice-impacted populations, offering potential avenues for intervention to improve health outcomes among this disadvantaged population.

## Methods

We used baseline data from the JUSTice-Involved Individuals Cardiovascular Disease Epidemiology (JUSTICE, R01 HL137696) study; a prospective, observational cohort whose protocol has been previously described in extensive detail [16]. The JUSTICE study enrolled individuals who had been recently released from jail or prison, had a CVD risk factor, and returned to New Haven, Bridgeport, and Hartford, Connecticut. Baseline data included a detailed questionnaire, point-of-care testing, and health literacy measured with the Short Test of Functional Health Literacy in Adults (S-TOFHLA) tool [16].

Between January 2019 and November 2023, participants were recruited based on time since release (within 90 days) and the presence of any of the following CVD risk factors: hypertension, diabetes, obesity, or hyperlipidemia. Potential participants were excluded if they had a known terminal illness or serious mental illness, or planned to relocate within 12 months, to mitigate attrition over the 12-month follow-up period. At the baseline visit, all participants provided written informed consent, and a trained research assistant administered a baseline questionnaire, gathering sociodemographic, clinical, psychosocial, and behavioral information, as well as exposure to criminal justice policies. CVD risk factors were objectively measured using point-of-care testing [16, 17]. The Yale University Human Investigation Committee (HIC#2000022213) and the Office of Human Research Protections approved this study.

### Study Measures

To assess health literacy, we chose the S-TOFHLA measure as it aligns with the Department of Health and Human Services definition of health literacy [18]. The S-TOFHLA assesses literacy using a 36-item questionnaire, with scores reported on a scale of 0 to 36, where scores of 0-16 indicate inadequate functional health literacy, 17-22 indicate marginal functional health literacy, and 23-36 indicate adequate functional health literacy [19]. For this study, we dichotomized the variable into inadequate health literacy (0-22) and adequate health literacy (23-36) to align with prior literature examining clinically meaningful differences in functional health literacy [20]. Of the 471 participants who were enrolled in the study, 439 had complete data on health literacy and formed the basis of our analysis. Analysis of participants with missing data on health literacy suggests that data were mostly missing at random (Table S1).

Our main outcome was cardiovascular health as measured by the American Heart Association’s Life’s Essential Eight (LE8) [21]. The LE8 score (0-100 scale) represents the average of eight individual components, comprising four health behaviors (diet, physical activity, sleep, and smoking) and four health factors (blood pressure, cholesterol, blood glucose, and body mass index) influencing cardiovascular health. The LE8 was selected as it provides a comprehensive, multidimensional measure of cardiovascular health.

In the general population, scores on the LE8 scale are interpreted as follows: 0 to 49 corresponds to low cardiovascular health, 50 to 79 indicates moderate health, and 80 to 100 reflects high cardiovascular health. An optimal LE8 component score of 100 represents an individual with blood pressure (<120 mmHg / <80 mmHg), blood glucose (HbA1c <5.7%), BMI (<25 kg/m^2^), non-high-density lipoprotein cholesterol (<130 mg/dL), no smoking history, diet score (8.0-10.0), and average sleep (7-9 hours/night), and physical activity (>150 minutes/week). Our team has previously published the scoring guide for each independent component in this cohort [17].

### Covariates

Baseline sociodemographic characteristics, including age, self-reported gender, race and ethnicity (Non-Hispanic White, Non-Hispanic Black, Hispanic, Other [composed of Native American, Asian, and Hawaiian]), education (Less than high school, GED, high school grad, some college/technical, College grad/grad school), marital status (single, married, widowed/separated/divorced), and income level (no income, less than $500, $500 to $999, $1000 and above, Unknown), were considered. We also included structural factors known to be linked to healthcare outcomes in this population, such as housing status (homeless, transitional home/halfway house, living with family/friends, and rent/own) and incarceration-related factors (times incarcerated, incarceration facility type, saw a Department of Corrections (DOC) provider while incarcerated, and time spent in solitary confinement) [22]. Health-related variables included diabetes, hyperlipidemia, hypertension, and drug and alcohol dependence, and were obtained from participant self-report of prior chronic disease diagnoses. Finally, psychosocial factors included self-efficacy scores, as measured by the General Self-Efficacy Scale [23].

### Analytical Approach

We determined the proportion of participants with adequate and inadequate health literacy based on S-TOFHLA scores. We then explored sociodemographic, structural, psychosocial, carceral, and clinical factors associated with health literacy using bivariate analyses to inform covariate selection for multivariable models. We used non-parametric tests, including the Wilcoxon rank sum test for continuous variables and Fisher’s exact or Pearson’s Chi-squared tests for categorical variables, consistent with non-normal distributions and small cell counts.

Next, we examined bivariate associations between health literacy and each LE8 component score. To further explore the relationship between health literacy and LE8 components, we conducted multivariable-adjusted linear regression analyses, treating LE8 components as quasi-continuous outcomes. Covariates associated with health literacy in bivariate analyses were included to account for potential confounding, alongside age, which was included a priori based on established associations with both health literacy and cardiovascular health [24, 25].

Statistical significance for all analyses was defined as 2-sided alpha=0.05. Data analysis was performed in RStudio (Version 2024.04.2+764) using the tidyverse, gtsummary, and broom packages.

## Results

Of the 439 participants with complete health literacy data, the mean age was 45 years (SD = 11); the majority were male (90%) and non-White (48% non-Hispanic Black, 27% Hispanic).

Educational attainment was low overall, with 23% of participants not completing high school and only 3.2% reporting a college degree or higher. Nearly four in five participants (78%) were living in transitional housing, while 4.1% reported being homeless at the first study visit. Over half (56%) reported three or more comorbid conditions, with hypertension being the most prevalent diagnosis (78%). Most participants (89%) had been incarcerated more than one time in their lifetime (Table 1).

**Table 1.**
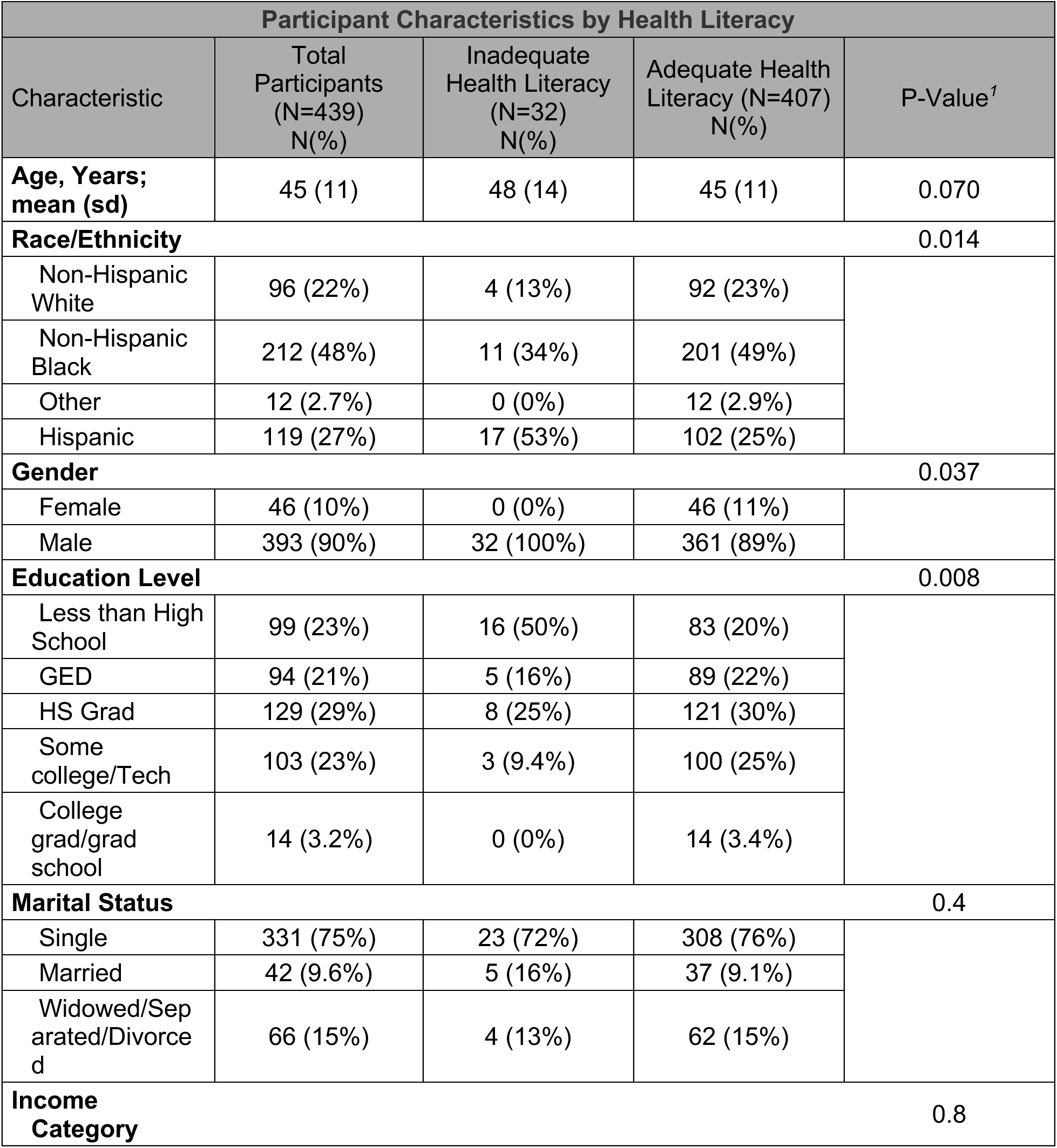
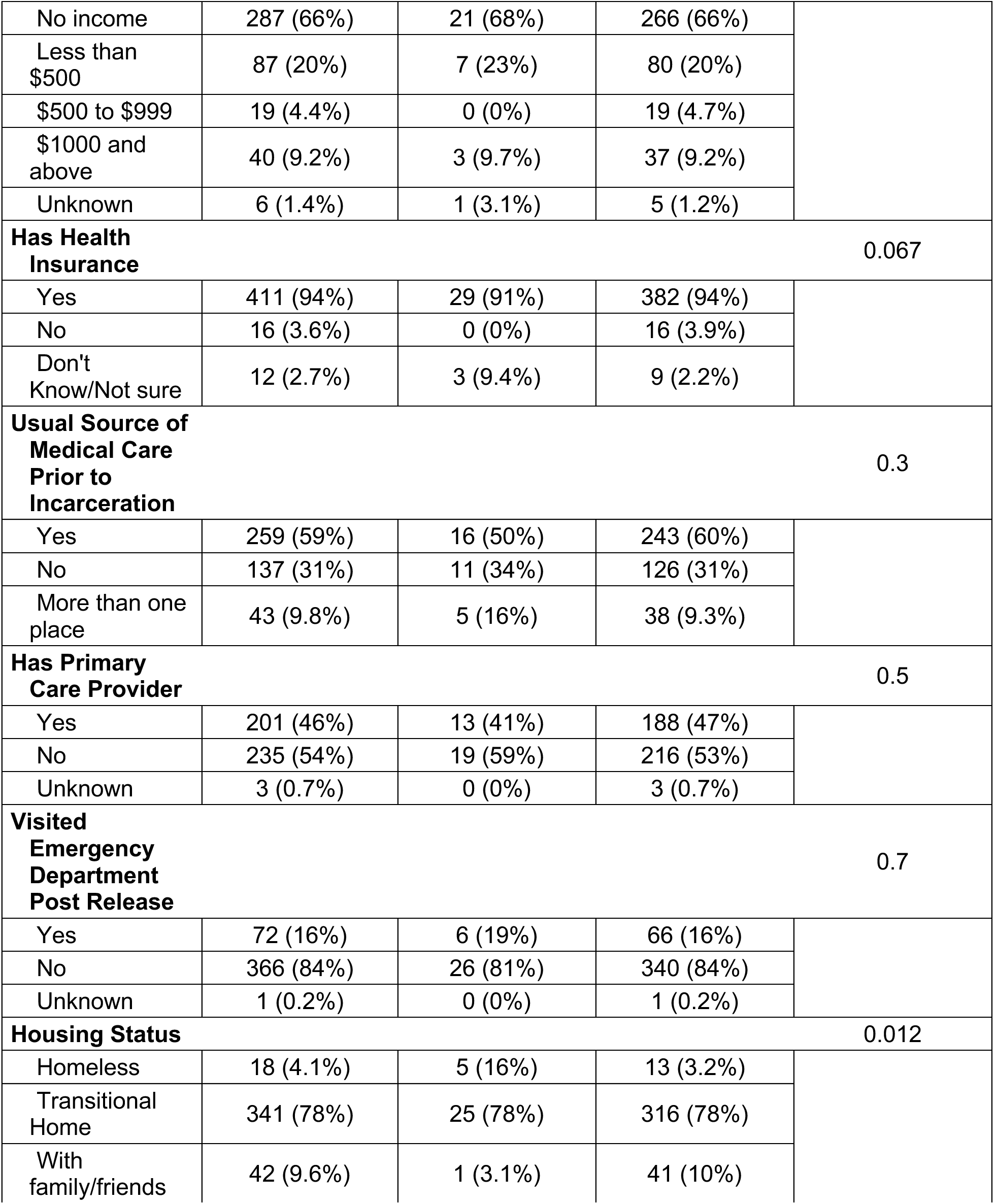
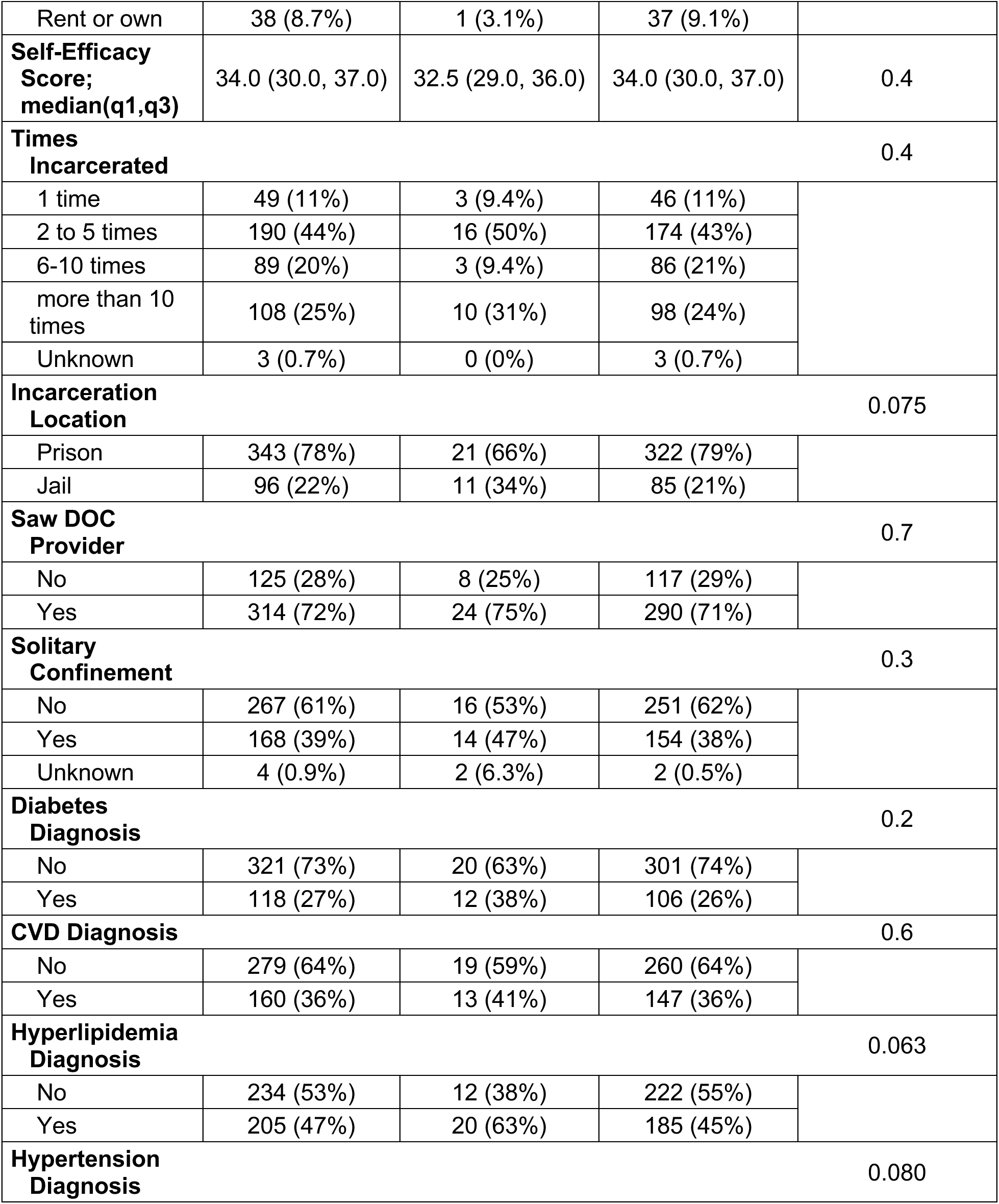
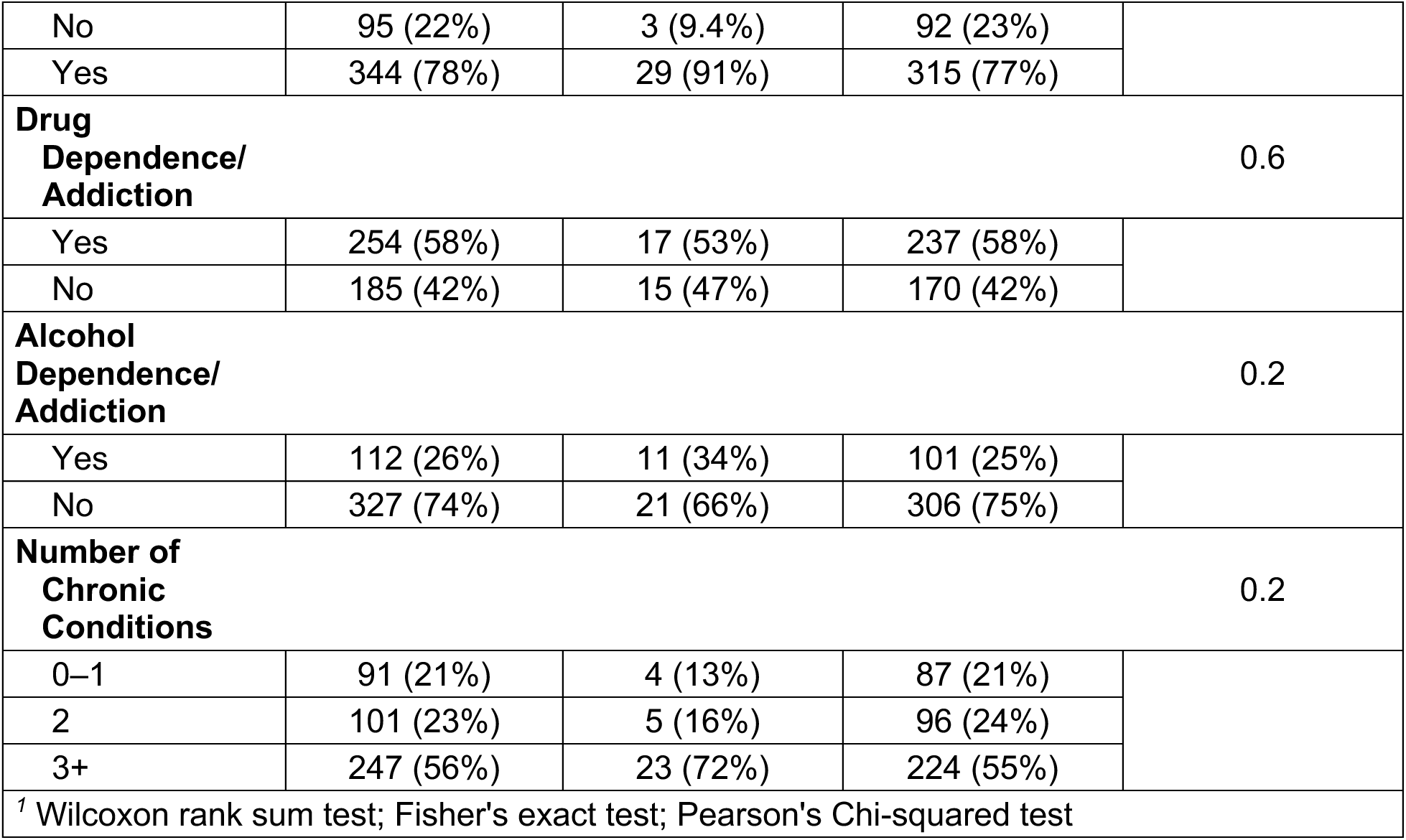
Participant Characteristics by Health Literacy Status. Baseline sociodemographic, incarceration-related, healthcare access, and clinical characteristics stratified by health literacy status (inadequate vs. adequate). Values are presented as mean (SD) for continuous variables, n (%) for categorical variables, and median (IQR) for self-efficacy scores. P-values were calculated using Wilcoxon rank-sum tests for continuous variables and Pearson’s chi-squared or Fisher’s exact tests for categorical variables, as appropriate.

Thirty-two participants (7%) had inadequate health literacy. Compared to those with adequate health literacy, participants with inadequate health literacy were significantly more likely to be Hispanic (53% vs. 25%, p *=* 0.014), male (100% vs. 89%, p = 0.037), and to have not completed high school (50% vs. 20%, p = 0.008). Those with inadequate health literacy were also more likely to be homeless (16% vs. 3.2%, p = 0.012) compared to those with adequate health literacy. Other characteristics, including income and number of prior incarcerations, were not significantly different by health literacy status (Table 1).

In bivariate analyses, individuals with inadequate health literacy had significantly lower scores on two LE8 components: physical activity (19.4 vs. 32.2, p = 0.03) and blood glucose (63.1 vs. 76.8, p = 0.02). Participants with inadequate health literacy were found to have a higher LE8 diet score (55.5 vs. 43.8, p = 0.04). No significant differences were observed in total LE8 score, BMI, blood pressure, cholesterol, smoking, or sleep scores (Table 2).

**Table 2.** LE8 Component Scores by Health Literacy Status. Mean Life’s Essential 8 (LE8) total and component scores are presented by health literacy status (inadequate vs. adequate). Values represent mean scores with corresponding standard errors (SE). P-values reflect unadjusted comparisons between groups.

| LE8 Component Scores by Health Literacy Status |  |  |  |  |  |  |  |  |
| --- | --- | --- | --- | --- | --- | --- | --- | --- |
| LE8 Component | N | LE8 Score (Total) |  | Inadequate Health Literacy |  | Adequate Health Literacy |  | p-value |
|  |  | Mean | SE | Mean | SE | Mean | SE |  |
| Body Mass Index | 433 | 41.82 | 1.49 | 42.03 | 5.61 | 41.81 | 1.54 | 0.96 |
| Blood Pressure | 437 | 47.03 | 1.39 | 46.88 | 4.86 | 47.04 | 1.45 | 0.97 |
| Blood lipids (non-HDL cholesterol) | 424 | 84.29 | 1.20 | 88.00 | 4.76 | 84.01 | 1.24 | 0.26 |
| Blood glucose | 425 | 75.74 | 1.47 | 63.12 | 6.00 | 76.77 | 1.50 | 0.02 |
| Physical activity | 424 | 31.23 | 1.71 | 19.38 | 6.04 | 32.19 | 1.78 | 0.03 |
| Diet | 432 | 44.62 | 1.51 | 55.47 | 5.01 | 43.75 | 1.58 | 0.04 |
| Smoking | 437 | 24.37 | 1.63 | 20.97 | 6.15 | 24.63 | 1.70 | 0.43 |
| Sleep health | 438 | 60.55 | 1.37 | 65.31 | 6.24 | 60.17 | 1.39 | 0.22 |
| Total score | 439 | 51.08 | 0.56 | 50.07 | 2.60 | 51.16 | 0.57 | 0.37 |

In linear regression models adjusted for age, gender, race/ethnicity, education, and housing status, inadequate health literacy was significantly associated with an average 11.9-point lower LE8 glucose score when compared with adequate health literacy (β = −11.9, 95% CI: −22.9 to −0.8, p = 0.04), but the associations with physical activity (p = 0.24) and diet (p = 0.12) were no longer significant (Figure 1).

**Figure 1.**
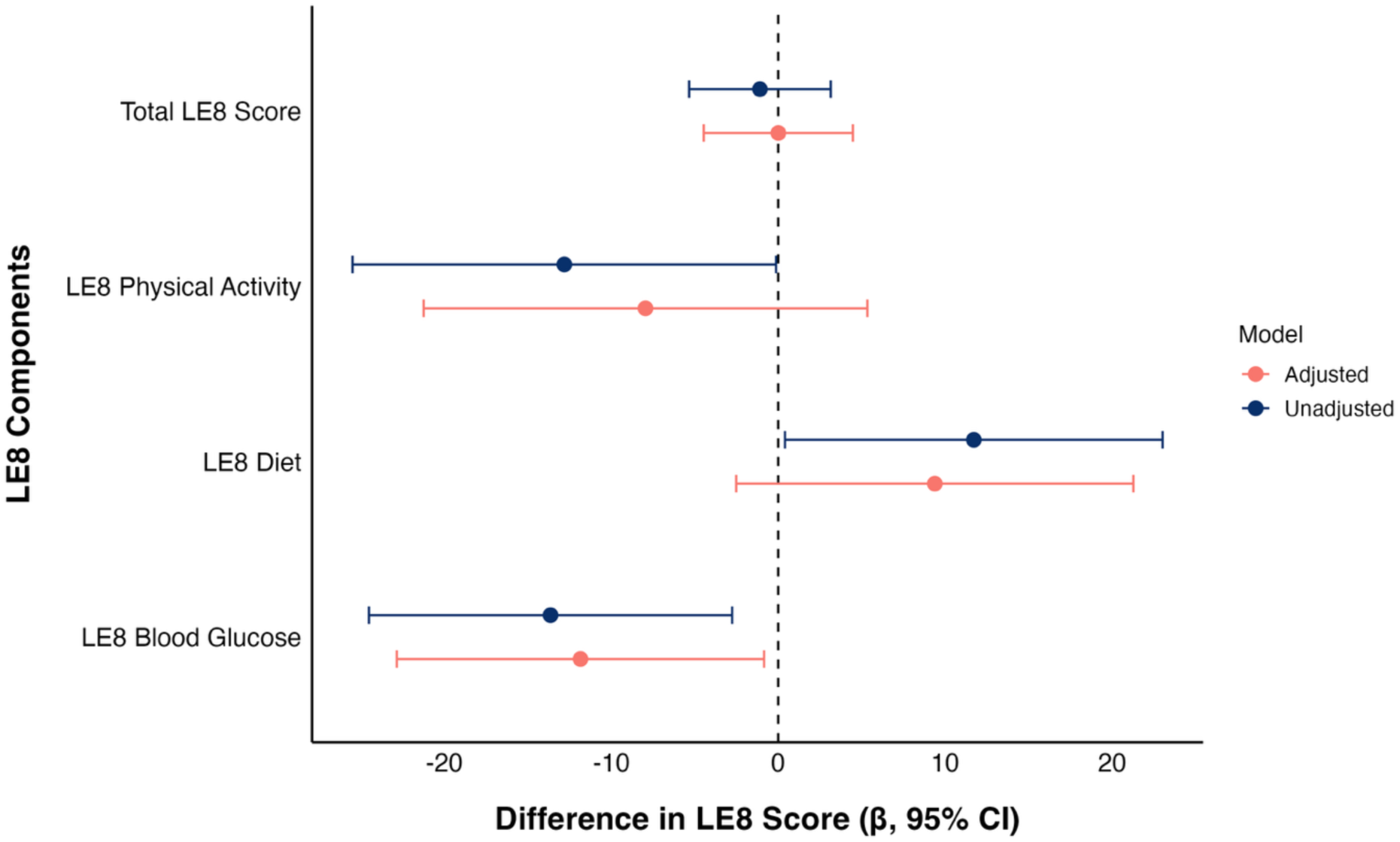
Association Between Inadequate Health Literacy and Life’s Essential 8 (LE8) Scores. Forest plot displaying unadjusted and adjusted associations between health literacy and total, as well as component, LE8 scores (physical activity, diet, and blood glucose). Points represent β estimates from linear regression models comparing participants with inadequate health literacy to the reference group, participants with adequate health literacy, and horizontal lines indicate 95% confidence intervals. The adjusted model accounts for age, gender, race/ethnicity, education level, and housing status. The dashed vertical line at zero represents no difference in LE8 score between participants with inadquate versus adequate health literacy.

## Discussion

In this study of justice-involved individuals recently released from incarceration, we found that 7% of participants had inadequate health literacy. While our findings on participant characteristics associated with health literacy are consistent with prior research highlighting disparities in literacy among formerly incarcerated individuals, the prevalence observed in our sample is notably lower than estimates reported in similar populations [6, 19, 26, 27]. This difference likely reflects a key feature of our study cohort. All participants were enrolled based on the presence of at least one CVD risk factor (hypertension, diabetes, obesity, or hyperlipidemia), with the majority reporting multiple chronic conditions. As a result, participants may have had more frequent engagement with the healthcare system than the general target population of formerly incarcerated persons, potentially leading to the observed higher rates of primary care before incarceration among those with adequate health literacy in our sample.

Consistent with these trends, a continuous patient-provider relationship has been associated with better health literacy, lessened treatment burden, and improved self-management of chronic conditions [28–30]. Additionally, health literacy was assessed using the S-TOFHLA, which emphasizes reading comprehension and applies less stringent thresholds for inadequate literacy compared to other instruments, such as the Newest Vital Sign [31, 32]. As a result, the S-TOFHLA may underestimate the prevalence of limited health literacy.

Health literacy was significantly associated with race, gender, educational attainment, and housing status in our sample. Consistent with past studies, we observed that inadequate health literacy was more common among individuals with lower educational attainment and those experiencing homelessness [33, 34]. We also found that health literacy was significantly associated with several components of cardiovascular health as measured by Life’s Essential 8 scores. In bivariate analyses, individuals with inadequate health literacy had significantly lower LE8 blood glucose scores than those with adequate health literacy, indicating that inadequate health literacy is significantly associated with poor glycemic control and high clinical HbA1c levels. Notably, the association between health literacy and blood glucose remained significant after adjusting for significant sociodemographic covariates. Blood glucose management relies heavily on understanding medication instructions, monitoring laboratory values, and navigating ongoing clinical care, underscoring the important role that health literacy plays in managing cardiovascular disease for this population [35–37].

In unadjusted analyses, inadequate health literacy was also negatively associated with the physical activity LE8 score but positively associated with participant diet scores. However, after adjusting for significant sociodemographic covariates, both associations were attenuated and no longer statistically significant. In our sample, the relationship between health literacy and physical activity was largely explained by educational attainment, which is strongly correlated with health literacy and shapes access to resources, opportunities, and environments that facilitate regular physical activity [2, 4, 38, 39]. Similarly, adjustment for age mitigated the association between health literacy and diet, which may reflect age-related differences in dietary patterns and food preferences [40, 41]. These findings suggest that while health literacy is an important determinant of cardiovascular health behaviors, its influence is also deeply shaped by broader social and demographic factors that influence individuals’ ability to access, understand, and utilize health information.

As a leading cause of death among incarcerated individuals, mitigating CVD risk is a critical goal for improving health outcomes for justice-involved populations [10]. Our findings highlight the important role that health literacy may play as a point of intervention. Within carceral facilities, individuals are typically highly restricted from monitoring their disease progression or self-medicating, undermining self-efficacy in chronic disease management. Interventions designed to support the health of incarcerated individuals should therefore consider the implementation of targeted educational services for patient empowerment. These efforts could include expanded access to self-monitoring blood glucose meters, which have shown promising results for those with progressive diabetes in prior initiatives by the Federal Bureau of Prisons [42]. Other efforts could include enhanced medical transparency from correctional health services, support groups led by health educators, and increased support for continuity of care post-release [43, 44].

This study is limited by its cross-sectional design that prevents conclusions about causality between health literacy and CVD-related health measurements. While LE8 scores were treated as continuous variables for regression modeling, their ordinal nature may limit the reliability of our estimates. Furthermore, the sample was drawn from individuals returning from a single state department of corrections to three study sites, limiting generalizability.

Future research is needed to examine how health literacy and CVD risk evolve in the reentry period, particularly with the provision of structured support. As efforts to improve carceral health outcomes continue, integrating health literacy into chronic disease prevention frameworks represents a powerful opportunity to reduce the disproportionate burden of cardiovascular disease in individuals involved in the criminal justice system.

## Conclusion

Our findings indicate that inadequate health literacy is associated with lower cardiovascular health, particularly in the domain of glycemic control, among individuals recently released from incarceration. Given the high burden of poor cardiovascular health among justice-involved populations and the rapid transition to self-directed care upon release, addressing health literacy is a critical and actionable pathway to improving health outcomes.

## Data Availability

The data that support the findings of this study are available from the senior author upon reasonable request.

## Acknowledgements

We thank the members of the study team who obtained informed consent and collected participant data. We also appreciate the outreach efforts of our community partners in identifying potential participants, and we acknowledge the collaboration of Intercommunity Health Care.

## Author contributions

BB involved in methodology, data curation, formal analysis, writing-original draft preparation, and writing-reviewing and editing. JAA involved in methodology, data curation, formal analysis, writing-reviewing and editing, and supervision. LBP involved in conceptualization, writing-reviewing and editing. EAW involved in conceptualization, methodology, funding acquisition, writing-reviewing and editing.

The analyses, interpretations, and conclusions presented here are solely the responsibility of the authors. They do not necessarily reflect the views of the National Heart, Lung, and Blood Institute, the U.S. Department of Health and Human Services, or any of their affiliates.

---

**Table S1.** Participant Characteristics by Health Literacy Missing Status. Baseline sociodemographic, incarceration-related, healthcare access, and clinical characteristics comparing participants with complete health literacy data (n = 439) to those with missing health literacy data (n = 32). Values are presented as mean (SD) for continuous variables, n (%) for categorical variables, and median (IQR) for self-efficacy scores. P-values were calculated using Wilcoxon rank-sum tests for continuous variables and Pearson’s chi-squared or Fisher’s exact tests for categorical variables, as appropriate. Participants with missing health literacy data were largely similar to those with complete data across sociodemographic characteristics, incarceration history, healthcare access, and most clinical conditions. A higher prevalence of diabetes (44% vs. 27%, p = 0.040) and differences in number of chronic conditions (p = 0.025) were observed among participants with missing data. Overall, the small proportion of missing observations (7%) and minimal differences across measured characteristics suggest limited evidence of systematic selection bias.

| Participant Characteristics by Missing Status |  |  |  |  |
| --- | --- | --- | --- | --- |
| Characteristic | Total Participants<br>(N=471)<br>N(%) | Included Participants<br>(N=439)<br>N(%) | Missing Participants<br>(N=32)<br>N(%) | P-Value<sup>1</sup> |
| Age, Years;<br>mean (sd) | 45 (11) | 45 (11) | 47 (10) | 0.2 |
| Race/Ethnicity |  |  |  | 0.8 |
| Non-Hispanic White | 101 (21%) | 96 (22%) | 5 (16%) |  |
| Non-Hispanic Black | 229 (49%) | 212 (48%) | 17 (53%) |  |
| Other | 13 (2.8%) | 12 (2.7%) | 1 (3.1%) |  |
| Hispanic | 128 (27%) | 119 (27%) | 9 (28%) |  |
| Gender |  |  |  | 0.4 |
| Female | 51 (11%) | 46 (10%) | 5 (16%) |  |
| Male | 420 (89%) | 393 (90%) | 27 (84%) |  |
| Education Level |  |  |  | 0.4 |
| Less than High School | 109 (23%) | 99 (23%) | 10 (31%) |  |
| GED | 97 (21%) | 94 (21%) | 3 (9.4%) |  |
| HS Grad | 138 (29%) | 129 (29%) | 9 (28%) |  |
| Some college/Tech | 112 (24%) | 103 (23%) | 9 (28%) |  |
| College grad/grad school | 15 (3.2%) | 14 (3.2%) | 1 (3.1%) |  |
| Marital Status |  |  |  | 0.8 |
| Single | 355 (75%) | 331 (75%) | 24 (75%) |  |
| Married | 44 (9.3%) | 42 (9.6%) | 2 (6.3%) |  |
| Widowed/Separated/Divorced | 72 (15%) | 66 (15%) | 6 (19%) |  |
| Income Category |  |  |  | 0.9 |
| No income | 309 (66%) | 287 (66%) | 22 (69%) |  |
| Less than \$500 | 92 (20%) | 87 (20%) | 5 (16%) | |
| \$500 to \$999 | 21 (4.5%) | 19 (4.4%) | 2 (6.3%) | |
| \$1000 and above | 43 (9.2%) | 40 (9.2%) | 3 (9.4%) | |
| Unknown | 6 | 6 | 0 |  |
| Has Health Insurance |  |  |  | 0.4 |
| Yes | 440 (93%) | 411 (94%) | 29 (91%) |  |
| No | 18 (3.8%) | 16 (3.6%) | 2 (6.3%) |  |
| Characteristic | Total Participants<br>(N=471)<br>N(%) | Included Participants<br>(N=439)<br>N(%) | Missing Participants<br>(N=32)<br>N(%) | P-Value<sup>1</sup> |
| Don't Know/Not sure | 13 (2.8%) | 12 (2.7%) | 1 (3.1%) |  |
| **Usual Source of Medical Care Prior to Incarceration** |  |  |  | 0.2 |
| Yes | 283 (60%) | 259 (59%) | 24 (75%) |  |
| No | 144 (31%) | 137 (31%) | 7 (22%) |  |
| More than one place | 44 (9.3%) | 43 (9.8%) | 1 (3.1%) |  |
| **Has Primary Care Provider** |  |  |  | 0.6 |
| Yes | 217 (46%) | 201 (46%) | 16 (52%) |  |
| No | 250 (54%) | 235 (54%) | 15 (48%) |  |
| Unknown | 4 | 3 | 1 |  |
| **Visited Emergency Department Post Release** |  |  |  | 0.091 |
| Yes | 81 (17%) | 72 (16%) | 9 (28%) |  |
| No | 389 (83%) | 366 (84%) | 23 (72%) |  |
| Unknown | 1 | 1 | 0 |  |
| **Housing Status** |  |  |  | 0.7 |
| Homeless | 19 (4.0%) | 18 (4.1%) | 1 (3.1%) |  |
| Transitional Home | 368 (78%) | 341 (78%) | 27 (84%) |  |
| With family/friends | 43 (9.1%) | 42 (9.6%) | 1 (3.1%) |  |
| Rent or own | 41 (8.7%) | 38 (8.7%) | 3 (9.4%) |  |
| **Self-Efficacy Score; median(q1,q3)** | 34.0 (30.0, 37.0) | 34.0 (30.0, 37.0) | 36.0 (31.0, 38.5) | 0.070 |
| **Times Incarcerated** |  |  |  | 0.2 |
| 1 time | 50 (11%) | 49 (11%) | 1 (3.2%) |  |
| 2 to 5 times | 205 (44%) | 190 (44%) | 15 (48%) |  |
| 6-10 times | 99 (21%) | 89 (20%) | 10 (32%) |  |
| more than 10 times | 113 (24%) | 108 (25%) | 5 (16%) |  |
| Characteristic | Total Participants<br>(N=471)<br>N(%) | Included Participants<br>(N=439)<br>N(%) | Missing Participants<br>(N=32)<br>N(%) | P-Value<sup>1</sup> |
| Unknown | 4 | 3 | 1 |  |
| **Incarceration Location** |  |  |  | >0.9 |
| Prison | 368 (78%) | 343 (78%) | 25 (78%) |  |
| Jail | 103 (22%) | 96 (22%) | 7 (22%) |  |
| **Saw DOC Provider** |  |  |  | 0.5 |
| No | 136 (29%) | 125 (28%) | 11 (34%) |  |
| Yes | 335 (71%) | 314 (72%) | 21 (66%) |  |
| **Solitary Confinement** |  |  |  | 0.4 |
| No | 289 (62%) | 267 (61%) | 22 (69%) |  |
| Yes | 178 (38%) | 168 (39%) | 10 (31%) |  |
| Unknown | 4 | 4 | 0 |  |
| **Diabetes Diagnosis** |  |  |  | 0.040 |
| No | 339 (72%) | 321 (73%) | 18 (56%) |  |
| Yes | 132 (28%) | 118 (27%) | 14 (44%) |  |
| **CVD Diagnosis** |  |  |  | 0.4 |
| No | 297 (63%) | 279 (64%) | 18 (56%) |  |
| Yes | 174 (37%) | 160 (36%) | 14 (44%) |  |
| **Hyperlipidemia Diagnosis** |  |  |  | 0.7 |
| No | 252 (54%) | 234 (53%) | 18 (56%) |  |
| Yes | 219 (46%) | 205 (47%) | 14 (44%) |  |
| **Hypertension Diagnosis** |  |  |  | >0.9 |
| No | 102 (22%) | 95 (22%) | 7 (22%) |  |
| Yes | 369 (78%) | 344 (78%) | 25 (78%) |  |
| **Drug Dependence/Addiction** |  |  |  | 0.081 |
| Yes | 273 (58%) | 254 (58%) | 19 (59%) |  |
| No | 197 (42%) | 185 (42%) | 12 (38%) |  |
| Don't Know | 1 (0.2%) | 0 (0%) | 1 (3.1%) |  |
| **Alcohol Dependence/Addiction** |  |  |  | 0.14 |
| Characteristic | Total<br>Participants<br>(N=471)<br>N(%) | Included<br>Participants<br>(N=439)<br>N(%) | Missing<br>Participants<br>(N=32)<br>N(%) | P-Value<sup>1</sup> |
| Yes | 124 (26%) | 112 (26%) | 12 (38%) |  |
| No | 347 (74%) | 327 (74%) | 20 (63%) |  |
| Number of Chronic Conditions |  |  |  | 0.025 |
| 0–1 | 92 (20%) | 91 (21%) | 1 (3.1%) |  |
| 2 | 113 (24%) | 101 (23%) | 12 (38%) |  |
| 3+ | 266 (56%) | 247 (56%) | 19 (59%) |  |
| <sup>1</sup> Wilcoxon rank sum test; Fisher's exact test; Pearson's Chi-squared test |  |  |  |  |

